# Multidrug therapy with terbinafine and itraconazole is not superior to itraconazole alone in current epidemic of altered dermatophytosis in India: A randomized pragmatic trial

**DOI:** 10.1101/19003947

**Authors:** Sanjay Singh, Vinayak N Anchan, Usha Chandra, Radhika Raheja

**Affiliations:** Department of Dermatology and Venereology, Institute of Medical Sciences, Banaras Hindu University, Varanasi, India

**Keywords:** Dermatophytes, Dermatophytosis, Itraconazole, Terbinafine, Tinea

## Abstract

**Background:** Treatment responsiveness of dermatophytosis has decreased considerably in recent past in India. We compared effectiveness of oral terbinafine daily (Terb) (active control) versus itraconazole daily (Itra) versus terbinafine plus itraconazole daily (TI) versus terbinafine daily plus itraconazole pulse (TIp) in tinea corporis, tinea cruris and tinea faciei in a pragmatic randomized open trial.

**Methods:** Ninety-two microscopically confirmed patients were allocated to Terb (6 mg/kg/day), Itra (5 mg/kg/day), TI (terbinafine 6 mg/kg/day, itraconazole 5 mg/kg/day), or TIp (terbinafine 6 mg/kg/day, itraconazole 10 mg/kg/day for 1 week in 4 weeks) group by concealed block randomization and treated for 8 weeks or cure.

**Results:** Cure rates were similar at 4 weeks (P=0.768). At 8 weeks, 5 (21.7%), 18 (78.3%), 16 (69.6%), and 16 (69.6%) patients were cured in Terb, Itra, TI, and TIp groups, respectively. All experimental regimens (Itra, TI, TIp) were more effective than Terb (P≤0.0027). All experimental regimens had similar effectiveness (P≥0.738). Relapse rates 4 and 8 weeks after cure were similar (P=0.869 and 0.314, respectively). Number-needed-to-treat (NNT) was 2 for Itra, 3 for TI, and 3 for TIp.

**Conclusions:** Oral itraconazole given daily (NNT=2) is the most effective treatment and combining it with terbinafine does not increase effectiveness.

**One Sentence Summary:** Combination of oral terbinafine and itraconazole is not more effective than itraconazole alone in current epidemic of altered dermatophytosis in India.

## Introduction

Unprecedented changes in the epidemiology, clinical features and treatment responsiveness of dermatophytosis have been noted in recent past in India.^1-4^ Many patients present with extensive dermatophytosis (Figure 1a). Recent data show that terbinafine, once a highly effective drug, now has an abysmal cure rate of 30.6% in tinea corporis, tinea cruris and tinea faciei when given orally at a dose of 5 mg/kg/day for 4 weeks.^5^ Decreased effectiveness of terbinafine has been attributed to mutation in the squalene epoxidase gene.^6, 7^ Furthermore, there is general impression and unpublished data with us of decreased effectiveness of other antifungal drugs also in India. Importance of finding effective methods of treating tinea cannot be overemphasized.

**Figure 1.**
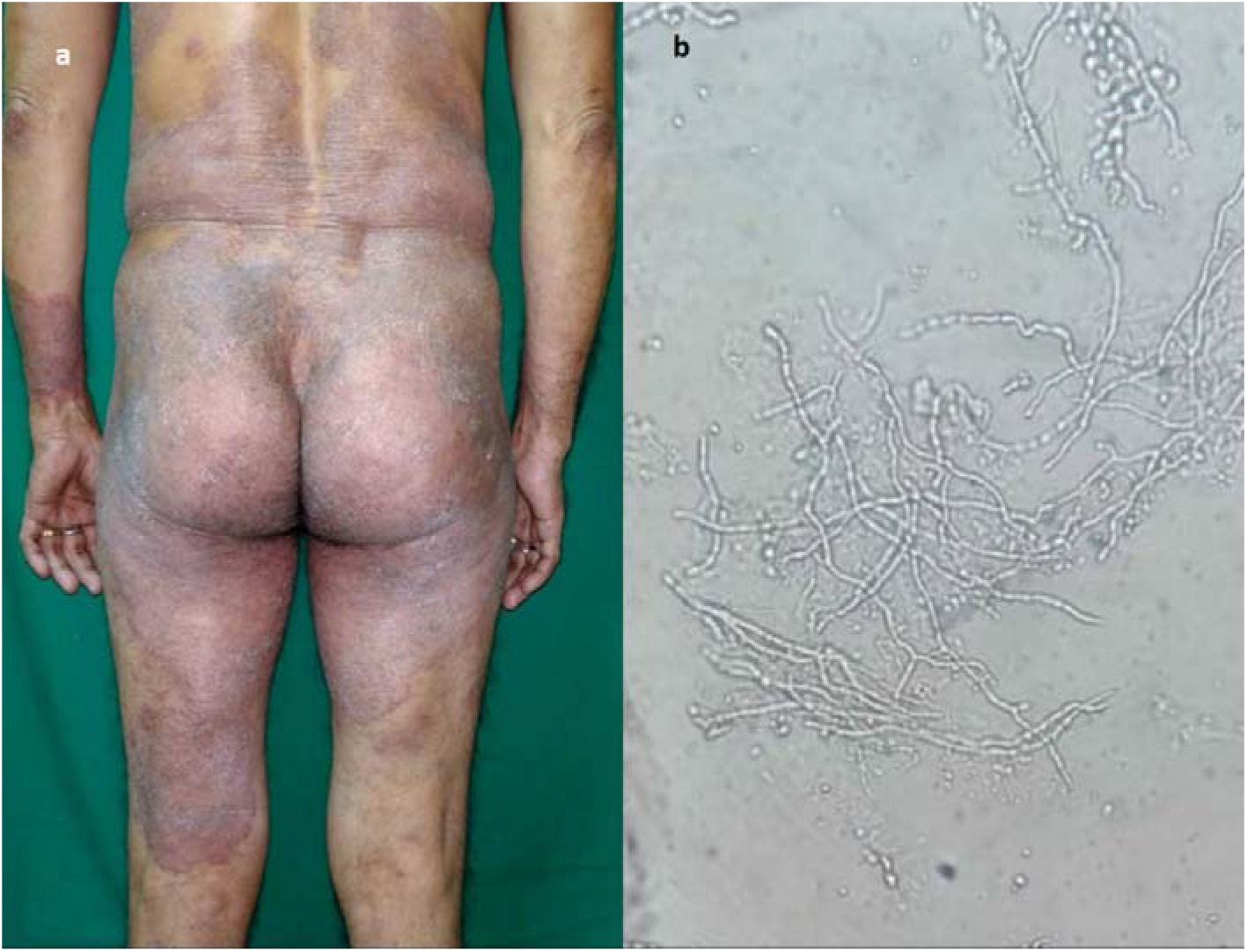
(a) A 55-year-old man with extensive tinea corporis, body surface area affected is 35%; (b) 20% potassium hydroxide microscopic (KOH) examination of skin scrapings showing septate, branching hyphae.

In the present study, we tested the hypothesis that using a regimen consisting of more than one oral antifungal drugs may produce better treatment outcome. We performed a four-arm pragmatic randomized open trial comparing effectiveness of oral terbinafine (Terb, active control) versus 3 experimental regimens, itraconazole daily (Itra) versus terbinafine plus itraconazole daily (TI) versus terbinafine daily plus itraconazole pulse (TIp).

## Materials and Methods

### Setting

The study (registered with Clinical Trials Registry-India, registration number CTRI/2018/09/015843) was conducted at a tertiary care centre after obtaining approval from the Institutional Ethics Committee.

### Sample size

A sample size of 23 patients in each group was determined using expected cure rates of 33.3% and 80% with the control and experimental regimens, respectively, with type I error rate of 0.05, type II error rate of 0.1 and dropout rate of 20%.^8^

### Selection criteria and enrolment

Patients with tinea corporis, tinea cruris or tinea faciei or a combination of these conditions were included in the trial. Inclusion criteria were (a) clinical suspicion of tinea corporis, tinea cruris or tinea faciei, (b) microscopic confirmation (potassium hydroxide [KOH] microscopy) (Figure 1b), and (c) age 4 to 80 years. Exclusion criteria were (a) presence of any other type(s) of tinea, e.g., onychomycosis, (b) pregnancy, (c) lactation, (d) inability to come for follow-up, (e) history of adverse reaction to terbinafine or itraconazole, (f) abnormal complete blood count (CBC), liver and renal function tests (LFT, RFT), (g) current treatment with drugs likely to cause interaction with terbinafine or itraconazole, (h) present history of renal, liver or ischemic heart disease, (i) presence of another skin disease at the site of tinea, (j) treatment with terbinafine or itraconazole in last one month, and (k) patients requiring antihistamines due to other skin disease. A witnessed, written and informed consent was given by the patients, or by a parent in case of minor patients. All female patients of child bearing age were advised to avoid pregnancy during treatment.

One thousand three hundred twenty six patients with suspected tinea corporis, tinea cruris or tinea faciei or a combination of these conditions attending dermatology outpatient department were assessed for eligibility. A total of 92 patients satisfied the selection criteria and were assigned to one of the four treatment groups by block randomization on the basis of random numbers generated online using Research Randomizer (https://www.randomizer.org/) (allocation ratio 1:1:1:1). Each treatment group comprised 23 patients (Figure 2). Patients were enrolled from October 2018 to December 2018. Allocation was concealed using sequentially numbered, sealed, opaque envelopes, which contained the allocation code written on a folded paper, the envelopes were opened after enrolment of the patients. Both random sequence generation and allocation concealment were done by a person unrelated to the trial.

**Figure 2.**
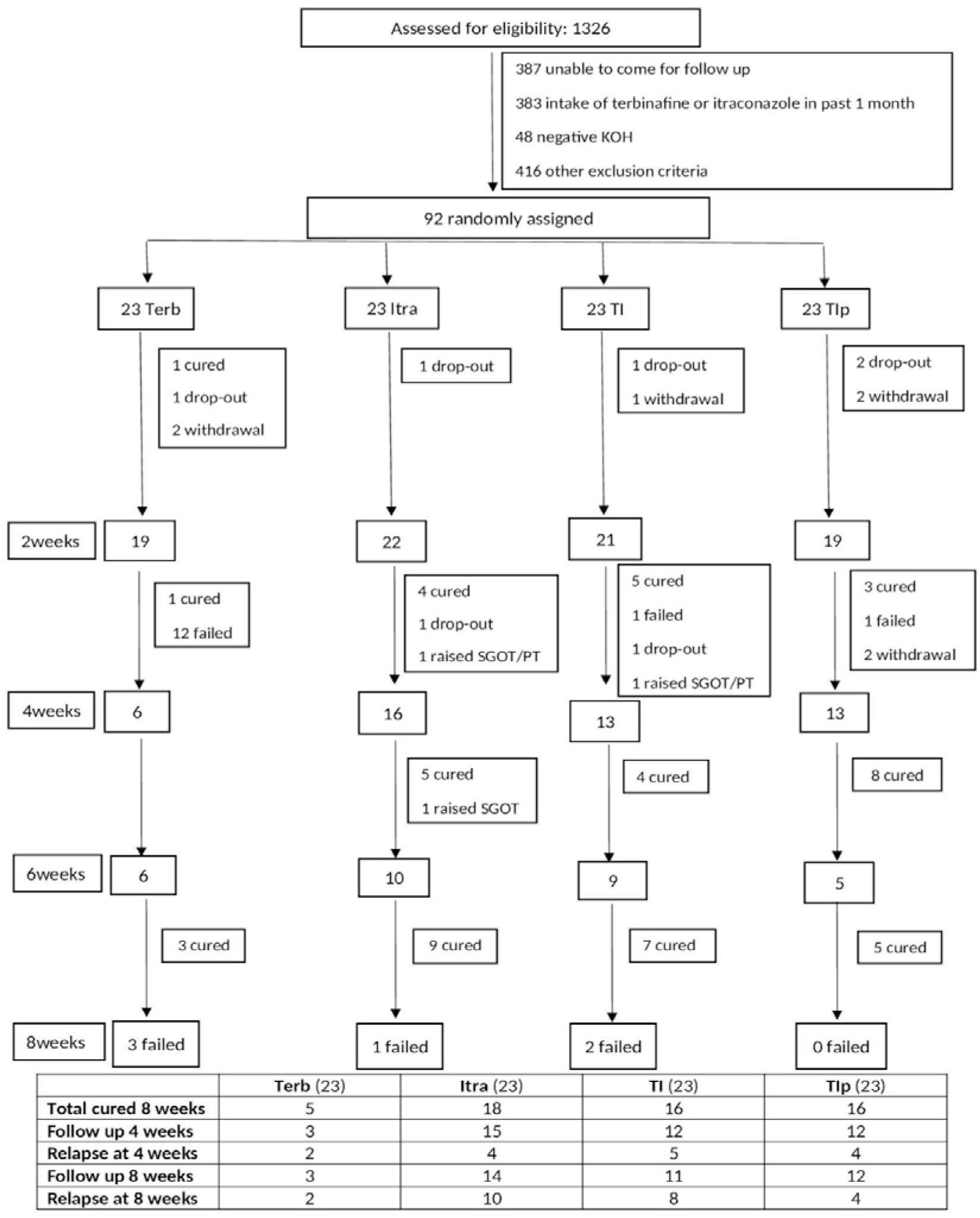
Flow diagram of the progress of the trial through its different phases. KOH, potassium hydroxide; SGOT, serum glutamic oxaloacetic transaminase levels; SGPT, serum glutamic pyruvic transaminase enzyme; Terb, Terbinafine daily; Itra, Itraconazole daily; TI, Terbinafine daily and Itraconazole daily; TIp, Terbinafine daily and Itraconazole pulse.

### Treatment

Doses given to the patients were as follows: Terb (terbinafine 6 mg/kg/day), Itra (itraconazole 5 mg/kg/day), TI (terbinafine 6 mg/kg/day, itraconazole 5 mg/kg/day), and TIp (terbinafine 6 mg/kg/day, itraconazole 10 mg/kg/day [with a maximum of 400 mg per day] for 1 week in 4 weeks). Patients received the treatment for 8 weeks or cure, whichever occurred earlier. Patients were advised against using any other treatment.

### Follow-up

Following data were recorded for each patient: body surface area (BSA) affected, treatment given, investigations CBC, LFT, and RFT at baseline, microscopy results, and follow-up data. In the terbinafine group, CBC and LFT were done every 2 weeks. In the patients who received itraconazole, following investigations were done every 2 weeks: CBC, LFT; and following every 4 weeks: RFT, fasting lipid profile, electrocardiogram, and urine routine and microscopy.

The most severe lesion was identified as the index lesion, from which scraping and KOH microscopy was performed at baseline and then at every subsequent visit. Patients were followed up at 2 weekly intervals up to a maximum of 8 weeks or cure, whichever occurred earlier.

### Measurement of treatment effect

Cure was defined as occurrence of both clinical cure (complete clearance of lesions) and mycological cure (negative KOH microscopy). Presence of post-inflammatory hyperpigmentation at the site of healed tinea was not considered a feature of tinea. Any of the following was considered treatment failure: (a) no improvement or worsening of the disease in 4 weeks in patient’s assessment, (b) presence of scaling and/or erythema at 8 weeks, and (c) positive microscopy at 8 weeks. Patients who were cured were asked whether they were fully satisfied with the treatment outcome or not. Patients in whom the treatment failed were treated outside the clinical trial as follows: terbinafine failures with itraconazole, itraconazole failures with terbinafine, and failures in the combination regimens with fluconazole.

### Measurement of compliance

Patients were asked to bring used strips of tablets and the strips in use at each follow up visit to assess compliance. Compliance was considered to be good if a patient had taken 80% or more of the prescribed number of tablets, and poor if less than 80% tablets were taken during treatment.

### Assessment for relapse

Those patients who were cured were assessed again 4 and 8 weeks after the cure to look for relapse of tinea (presence of erythema and scaling at the site of healed tinea).

### Outcome measures

Number of patients cured at 8 weeks with the different regimens was the primary outcome measure. Secondary outcome measures included number of patients cured at 4 weeks, relapse rate, and frequency of adverse events. In addition, severity of itching was noted on a 0 (no itching) to 10 (most severe itching) at baseline and subsequent follow-ups.

### Statistical analysis

For the baseline variables, mean and standard deviation (SD) or median and interquartile range (IQR, IQ1-IQ3) were calculated depending on the distribution of data. Cure rates, relapse rates and compliance were compared by Fisher exact test. P values less than 0.05 were considered significant. All P vales are two-tailed. Absolute risk reduction (ARR) of treatment failure versus the control (terbinafine daily) and number needed to treat (NNT) were calculated. When appropriate, 95% confidence intervals (CI) were calculated. Intention to treat analysis was performed for the effectiveness data. Compliance data were analysed on per protocol basis. Denominator for calculating compliance was the total number of patients who were cured and those who came for follow ups up to 8 weeks but were not cured. Denominator for calculation of relapse rate was the number of patients who had achieved cure. For calculating relapse rate, those cured patients who could not be followed up were considered relapsed.

## Results

### Baseline characterises of the patients

Patients in the 4 groups were comparable at baseline with regard to age, duration of tinea, and body surface area affected (Table 1). Sixty patients had tinea corporis et cruris, 14 tinea corporis et cruris et faciei, 9 tinea corporis, 6 tinea cruris, 2 tinea fasciei, and 1 had tinea corporis et faciei. Four patients had received oral antifungal treatment in past (none received terbinafine or itraconazole in past 1 month). Seventeen, 19, 18, and 20 patients in the Terb, Itra, TI, and TIp groups, respectively, had applied some topical preparations in the past.

**Table 1.**
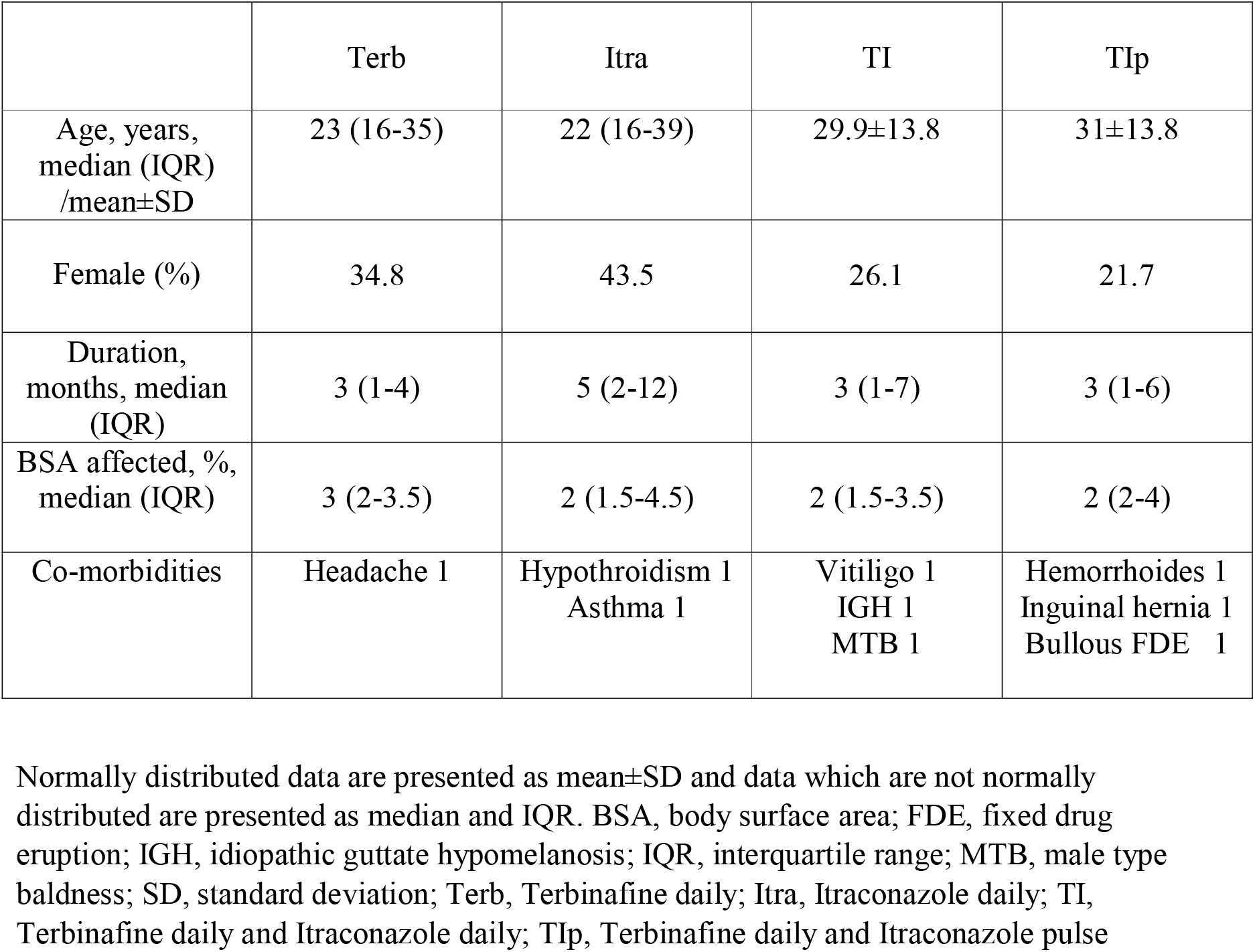
Baseline characteristics of the patients.

**Table 2.** Cure rates at 8 weeks and relapse rates with the four treatments.

| **Treatment** | **Patients cured,**<br>**n (% , 95% CI)** | **Patients who**<br>**relapsed within 4**<br>**weeks of cure,\* n**<br>**(%, 95% CI)** | **Patients who**<br>**relapsed within 8**<br>**weeks of cure,\* n**<br>**(%, 95% CI)** |
| --- | --- | --- | --- |
| Terbinafine | 5 (21.7, 9.2 to 42.3) | 2 (40, 11.6 to 77.1) | 2 (40, 11.6 to 77.1) |
| Itraconazole | 18 (78.3, 57.7 to 90.8) | 4 (22.2, 8.5 to 45.8) | 10 (55.6, 33.7 to 75.5) |
| Terbinafine+Itraconazole<br>daily | 16 (69.6, 48.9 to 84.6) | 5 (31.3, 13.9 to 55.9) | 8 (50, 28.0 to 72.0) |
| Terbinafine+Itraconazole<br>pulse | 16 (69.6, 48.9 to 84.6) | 4 (25, 9.7 to 50.0) | 4 (25, 9.7 to 50.0) |

\*includes those patients who could not be followed up to detect relapse; CI, confidence interval

### Cure rates at 4 weeks

At 4 weeks, 2 (8.7%), 4 (17.4%), 5 (21.7%), and 3 (13.0%) patients were cured in Terb, Itra, TI, and TIp groups, respectively. The cure rates were not significantly different (P=0.768) (Figure 2).

### Cure rates at 8 weeks

At 8 weeks, 5 (21.7%), 18 (78.3%), 16 (69.6%), and 16 (69.6%) patients were cured in Terb, Itra, TI, and TIp groups, respectively. Comparison of different pairs of treatment groups showed that all experimental regimens (Itra, TI, TIp) were more effective than Terb (P≤0.0027). Cure rates with the three experimental regimens were not significantly different (P≥0.738) (Figure 2, Table 3).

**Table 3.** Comparison of the cure rates at 8 weeks with the four treatments.

| **Treatment pairs** | **P** | **95% CI of difference** |
| --- | --- | --- |
| Terb vs Itra | 0.0003 | 28.0 to 73.6 |
| Terb vs TI | 0.0027 | 19.1 to 67.0 |
| Terb vs TIp | 0.0027 | 19.1 to 67.0 |
| Itra vs TI | 0.7381 | -16.3 to 32.4 |
| Itra vs TIp | 0.7381 | -16.3 to 32.4 |
| TI vs TIp | 1 | -25.3 to 25.3 |

Terb, Terbinafine daily; Itra, Itraconazole daily; TI, Terbinafine daily and Itraconazole daily; TIp, Terbinafine daily and Itraconazole pulse; CI, confidence interval

### Severity of itching

Severity of itching decreased in all treatment groups as the treatment progressed and itching was absent in all patients at the time of cure (Table 4).

**Table 4.** Severity of itching.

|  |  | Baseline | 4 weeks | 8 weeks |
| --- | --- | --- | --- | --- |
| All patients | Terb | 5.87±2.36 | 5 (2-7) | 0.5 (0-0) |
|  | Itra | 5.74±2.42 | 0 (0-1.75) | 0 (0-0) |
|  | TI | 6.43±2.59 | 0 (0-2.5) | 0 (0-0) |
|  | TIp | 6.22±2.19 | 2 (1-5) | 0 (0-0) |
| Patients cured upto 4 weeks | Terb | 8.5 (8.25-8.75) | 0±0 |  |
|  | Itra | 5 (4-5.75) | 0±0 |  |
|  | TI | 8 (8-8) | 0±0 |  |
|  | TIp | 8 (6.5-8.5) | 0±0 |  |
| Patients cured upto 8 weeks (excluding those cured upto 4 weeks) | Terb | 5 (5-6.5) | 5 (5-5) | 0±0 |
|  | Itra | 5 (5-8) | 3 (1.75-4.75) | 0±0 |
|  | TI | 6 (5-8.5) | 4 (2.75-5) | 0±0 |
|  | TIp | 5 (5-8) | 2 (2-5) | 0±0 |

Normally distributed data are presented as mean±SD and data which are not normally distributed are presented as median and IQR. Terb, Terbinafine daily; Itra, Itraconazole daily; TI, Terbinafine daily and Itraconazole daily; TIp, Terbinafine daily and Itraconazole pulse

### Relapse rate at 4 and 8 weeks

Two out of 5 cured patients (40.0%), 4 of 18 (22.2%), 5 of 16 (31.3%), and 4 of 16 (25.0%) patients relapsed within 4 weeks of cure in Terb, Itra, TI, and TIp groups, respectively. Relapse rates were not significantly different (P=0.869) (Figure 2, Table 2).

By 8 weeks, 2 (40.0%), 10 (55.6%), 8 (50.0%), and 4 (25.0%) patients had relapsed in Terb, Itra, TI, and TIp groups, respectively. Relapse rates at 8 weeks were not significantly different (P=0.314) (Figure 2, Table 2).

### Compliance

Twenty out of 20 patients, 20 of 22, 19 of 21, and 18 of 19 patients had good compliance in Terb, Itra, TI, and TIp groups, respectively. The difference was not significant (P= 0.714).

### Patients’ satisfaction with treatment outcome

All patients, who were cured, were fully satisfied with the outcome.

### Number needed to treat

Number needed to treat (NNT) for the experimental groups were as follows: 2 for Itra, 3 for TI, and 3 for TIp (Table 5).

**Table 5.** Number needed to treat and absolute risk reduction with the experimental treatments.

| **Treatment** | **NNT** | **ARR, %** | **95% CI of ARR, %** |
| --- | --- | --- | --- |
| Itra | 2 | 56.5 | 32.7 to 80.4 |
| TI | 3 | 47.8 | 22.6 to 73.1 |
| TIp | 3 | 47.8 | 22.6 to 73.1 |

Absolute risk reduction of treatment failure versus the control (terbinafine daily, Terb).
Itra, Itraconazole daily; TI, Terbinafine daily and Itraconazole daily; TIp, Terbinafine daily and Itraconazole pulse. NNT, number needed to treat; ARR, absolute risk reduction; CI, confidence interval

### Adverse events

In the Itra group, one patient at 4 weeks had raised serum glutamic pyruvic transaminase enzyme (SGPT) and serum glutamic oxaloacetic transaminase levels (SGOT) (both >100 u/l), and one patient at 6 weeks had raised SGOT (>100 u/l). One patient in TI group had raised SGPT and SGOT (both >100 u/l) at 4 weeks. Treatment was stopped in these 3 patients. Test results became normal in subsequent investigations performed after 2 weeks. No other adverse events were detected on investigations and none were reported by the patients.

## Discussion

Present study was conducted in view of unprecedented changes in the epidemiology, clinical features and treatment responsiveness of tinea infections in recent past in India.^1-4^ Recent data show that terbinafine, once a highly effective drug, now has abysmal cure rate in tinea corporis, tinea cruris and tinea faciei.^5^ These results confirmed the general impression among Indian dermatologists that terbinafine is fast losing its effectiveness. There is also a general impression, and we have unpublished data, that effectiveness of other antifungal drugs has also declined.

Terbinafine has been considered to be the oral antifungal drug of choice for dermatophytosis due to its high cure rates, fungicidal action, minimal drug interactions and a general lack of serious adverse effects. However, evidence shows that this is no longer the case in the current epidemic of altered dermatophytosis in India, which is characterized by altered morphology, extensiveness, abysmal responsiveness to terbinafine,^5^ and chronicity and frequent relapses within weeks of apparent cure.^9^ It is obviously important that new treatment methods are tested for their effectiveness in tinea infections in the current situation.

We tested the hypothesis that using a multidrug therapy regimen may produce better treatment outcome in tinea. Terbinafine and itraconazole were selected as the two antifungal agents for multidrug therapy because they have different mechanisms of action. Effectiveness of oral terbinafine daily (Terb) (active control) was compared with itraconazole daily (Itra), terbinafine plus itraconazole daily (TI), and terbinafine daily plus itraconazole pulse (TIp) in tinea corporis, tinea cruris and tinea faciei within a pragmatic randomized open design.

Cure rates at 4 weeks with the four treatments were not significantly different. Comparison of cure rates at 8 weeks with different pairs of treatment groups showed that all experimental regimens (Itra, TI, TIp) were more effective than Terb. Although cure rates with all experimental regimens were not significantly different at 8 weeks, number needed to treat (NNT) showed that itraconazole given alone was the most effective treatment.

NNT for the experimental group Itra (itrconazole alone) was 2, and that for TI and TIp each, it was 3; meaning thereby that, compared to Terb, 3 patients need to be treated with TI or TIp to achieve one additional cure, while only 2 patients need to be treated with Itra for the same effect. Thus, itraconazole was the most effective treatment out of the four treatments tested in the study.

Compliance and relapse rates were similar in the four groups. Of the 92 patients, only 3, who received itraconazole daily (one of them received terbinafine in addition), had reversible raised liver enzyme levels in the present study. However, the importance of closely monitoring liver enzymes cannot be overemphasized when treating patients with itraconazole or terbinafine.

A limitation of the present study is that it was not blinded.

Data presented suggest that effectiveness of oral terbinafine in tinea has declined further compared to the very recent data (30.6%^5^ versus 8.7% cure rate at 4 weeks in the present study). However, this can not be definitively commented upon because the present study was not designed to find out effectiveness of a single agent, which would require a larger sample size.

Data presented herein show that treatment with itraconazole alone given daily is the most effective treatment for patients with tinea corporis, tinea cruris and tinea faciei in the current epidemic of altered dermatophytosis in India, and that the addition of terbinafine to itraconazole regimen does not increase its effectiveness.

## Data Availability

Not available online.

